# Less is more: modelling the impact of species-targeted versus broadcast larviciding approaches for malaria control in rural settings

**DOI:** 10.64898/2026.03.04.26347561

**Authors:** Betwel J. Msugupakulya, Fredros O. Okumu, Anne L Wilson, Prashanth Selvaraj

## Abstract

**Background:** Larval source management (LSM) was once central to malaria control before insecticide-treated nets and indoor residual spraying dominated. Renewed interest in LSM raises questions about its effectiveness in rural Africa, where habitats are dispersed, and vector species contribute unequally, and whether species-targeted larviciding could offer greater gains than broadcast approaches.

**Methods:** This modelling study quantified the potential impact of larviciding in African settings where multiple vector species contribute unequally to malaria transmission. We modeled malaria transmission in southeastern Tanzania using agent-based simulations incorporating seasonal dynamics, insecticide resistance, and semi-field biolarvicide efficacy. Outcomes were entomological inoculation rate, malaria incidence in under-fives, and operational larviciding costs.

**Findings:** Large-scale deployment of biolarvicides with >1-week residual activity substantially reduced malaria transmission, with disproportionately greater gains when control efforts were preferentially focused on the dominant vector species, *Anopheles funestus*, compared to broadcast approaches treating both *An. funestus* and *An. arabiensis* habitats. In the absence of ITNs, a four-month fortnightly larviciding campaign targeting *An. funestus* at 80% coverage reduced EIR by 58% and incidence by ∼40%, versus ∼55% incidence and ∼70% EIR reductions under broadcast strategies; targeting *An. arabiensis* alone yielded ≤30% EIR and ≤13% incidence reductions. Starting with pre-existing 80% ITN coverage, *funestus*-targeted larviciding further reduced peak EIR by ∼70% and incidence by ∼77%, versus ∼90% and ∼85%, respectively, with broadcast strategies, suggesting broadcast larviciding provided limited additional reductions beyond those achieved by the *funestus*–targeted approach. At 40% ITN coverage, additional reductions were ∼62% of EIR and ∼46% in incidence (*funestus-*targeted) versus ∼76% and 63%, respectively (broadcast). The targeted campaigns preserved a 30–50% cost advantage while sustaining >50% dry-season transmission reductions. Finally, high-coverage (e.g., 80%) *funestus-*targeted larviciding campaigns achieved greater impacts than lower-coverage (e.g., 40-60%) targeting both species.

**Conclusions:** In settings where multiple vector species contribute unequally to malaria transmission, preferentially targeting larviciding against the dominant vector species can deliver substantial epidemiological impact, with greater resource efficiency than broadcast approaches targeting multiple vectors. In Tanzania, where *An. funestus* drives most transmission; concentrating larviciding efforts on its characteristic aquatic habitats may offer a scalable, low-cost complement to established tools such as ITNs.

## Background

Despite the gains brought by previous efforts, it is becoming clearer that the current malaria control interventions will not be enough to eliminate malaria or allow us to reach the ambitious goals set for malaria control by 2030 (WHO, 2015). Since 2015, the rate of progress against malaria has significantly reduced, a situation worsened in 2020 by the disruption of control during the COVID-19 pandemic [2]. The sustained effectiveness of insecticide-treated nets (ITNs), indoor residual spraying (IRS), and anti-malaria therapies is compromised by a combination of vector behavioral changes, increasing insecticide resistance, rising *Plasmodium* drug resistance, and climate change–driven shifts in vector ecology and malaria transmission [3–14]. Besides, the roles of different vector species in malaria transmission are now changing, likely due to the wide use of ITNs and IRS. For instance, *Anopheles funestus* is now the most important malaria vector in most areas across east and southern Africa [15]. Because of these ongoing challenges and the changing entomological landscape, vector control strategies must be revisited to ensure they are both effective and sustainable to further our progress toward the goals set.

Larval source management (LSM), which aims to target immature stages of mosquitoes (eggs, larvae, and pupae) in aquatic habitats, is now increasingly recognised as a potential intervention to supplement ITNs and IRS. LSM was historically an important component of malaria control efforts and was even used to eliminate certain disease vectors in some settings [16–19]. However, it was later deprioritized as malaria control efforts shifted predominantly toward adult mosquito control, driven in part by the availability of highly effective insecticides, including dichlorodiphenyltrichloroethane (DDT) [20,21]. In recent decades, successful LSM campaigns have also been demonstrated in several countries across Africa [22,23]. Yet, despite historical and recent evidence of its efficacy, LSM remains deprioritized across sub-Saharan Africa and is recommended only as a supplement to ITNs and IRS in settings where mosquito aquatic habitats are few, fixed, and findable [24].

LSM implementation is typically done as a broadcast intervention, which entails deploying one or more LSM techniques in all or a large proportion of detectable potential aquatic habitats of malaria vectors in a target area [25]. Targeting all aquatic habitats, particularly in rural areas where habitats are constantly changing, is likely to be beyond the resource capacities of most Sub-Saharan African countries.

Therefore, vector control interventions need to be tailored to the local needs to increase feasibility and cost-effectiveness, for example, only targeting productive habitats [26]. Although it is difficult to identify productive habitats in the field [27,28], a recent study in Burkina Faso demonstrated the high efficacy of targeting the most productive habitats based on remote sensing, which led to a 61% reduction in densities of malaria vectors compared to a 70% reduction using broadcast larviciding [23]. LSM can also be targeted to control vectors during the seasons when their populations are most vulnerable, typically during the dry season [29]. This requires a thorough understanding of the seasonal ecology of malaria vectors, including their refugia, where they can survive and persist until the rainy season returns. Targeted LSM can also be achieved by targeting habitats of a specific mosquito species that plays a major role in malaria transmission. This approach was used to great success in Malaysia when LSM was targeted at shaded habitats preferred by the mosquito species *An. umbrosus*, at the time termed ‘species sanitation’ [30,31]. For this strategy to be effective, the targeted vectors must have specific preferences for habitats with distinct characteristics that can be easily identified and managed.

Malaria transmission in Africa is dominated by four major malaria vector species, *An. gambiae, An. funestus, An. Arabiensis,* and *An. coluzzii*, alongside several other species, play a role in local settings [32]. Evidence suggests that in most settings across east and southern Africa, *An. funestus* is responsible for most of the malaria transmission [15]. Unfortunately, in several settings, this vector has increasingly shown higher resistance against the insecticides used in ITNs than other important vectors, likely contributing to its persistence despite the wide use of ITNs [3]. Studies on the aquatic ecology of this vector have demonstrated that *An. funestus* prefers unique larval habitats that can be few and fixed and potentially be targeted with larviciding [33]. Furthermore, *An. funestus* relies on these habitats during the dry season for proliferation, when it maintains transmission in the area, while the role of *An. arabiensis* is significantly low [4,29]. Despite these observations, there have been a few explorations of the potential impact of different larviciding strategies to inform an optimal larviciding strategy in the east and southern African settings, where *An. funestus* is the dominant malaria vector. In particular, there has not been any effort to assess whether a more limited strategy that preferentially targets the habitats of major malaria vectors would perform as well as a non-targeted (broadcast) approach.

Mathematical modelling can be used to explore questions in silico before large-scale implementation takes place and to inform the design of field trials. In this study, we combined agent-based malaria transmission modelling with experimental data on the residual efficacy of commercially available biolarvicides to quantify the potential impact of alternative larviciding strategies under realistic rural African settings where multiple vector species contribute unequally to malaria transmission. We used a calibrated individual-based model of malaria transmission in rural south-eastern Tanzania, where *An. funestus* is the dominant malaria vector species, but where *An. arabiensis* also remains widespread [4,34,35], to compare broadcast larviciding with strategies that preferentially target aquatic habitats of specific vector species. This approach allowed us to assess the epidemiological and operational trade-offs between species-targeted and non-targeted larviciding strategies across different levels of ITN coverage, intervention timing, and operational coverage.

## Methods

### Simulation framework

All simulations are performed with modelling software EMOD v2.20 [36], an individual-based mechanistic model of *Plasmodium falciparum* transmission. The model tracks individual mosquito vectors and human hosts through its three major components: Vectors are tracked through the vector life cycle and vector feeding cycle components, and humans are tracked through within-host parasite and immune dynamics. In the EMOD model, mosquito vectors pass through four stages: eggs, larvae, immature adult mosquitoes (that do not seek hosts), and mature adults (that seek hosts). Mature mosquito vectors produced from the vector life cycle enter the feeding cycle, which continues until the mosquito dies. The feeding cycle includes host-seeking, feeding, and ovipositing. The number of habitats each month for different vectors involved in malaria transmission was calibrated so that the resulting mosquito seasonality in the model was similar to that observed in field surveys conducted in rural south-eastern Tanzania [37]. The model also included a factor of habitat co-sharing, whereby 8% of habitats contained both *An. arabiensis* and *An. funestus* based on previous field data [37].

Indoor feeding fractions were assumed to be 63.1% for *An. arabiensis* and 78.2% for *An. funestus* based on a recent study in villages of south-eastern Tanzania [38]. To account for operational effectiveness, we incorporated the data on ITNs tested in experimental houses in northern Tanzania [39], with an initial killing effect of approximately 70%. We assumed that ITNs underwent exponential decay, leading to a decline in killing efficacy to 40% at the beginning of the second-year post-implementation. We also assumed the ability of ITNs to block biting mosquitoes was 95%, with exponential decay over four years. In addition, insecticide resistance to the insecticides used in bed nets was incorporated into the model, linking vector resistance to specific allele combinations within individual vectors. Thus, the phenotypic resistance was influenced by the expression of specific allele combinations within individual vectors.

Resistance was modeled as a factor reducing the effectiveness of insecticides used in ITNs, where we used resistance assay values as the probability of a resistant mosquito dying when encountering ITNs. Mosquitoes with co-dominant resistance alleles were assumed to display resistance levels approximately half that of those with homozygous resistance. *An. arabiensis* with a homozygous resistance allele was thus considered to exhibit pyrethroid resistance of 89% (mortality of 11%) and heterozygous *An. arabiensis* exhibits pyrethroid resistance of 44.5% (mortality of 55.5%), whereas *An. funestus* homozygous exhibited pyrethroid-resistant mosquitoes at 98% (mortality of 2%), and heterozygous *mosquitoes* exhibited pyrethroid resistance at 49% (mortality of 51%). These values are based on a bioassay conducted against wild mosquitoes collected in southeastern Tanzania (*Msugupakulya, Unpublished*).

The model incorporates a treatment-seeking intervention to simulate health-seeking behaviour and antimalarial treatment with artemether-lumefantrine, assuming that 80% of individuals have access to care. For uncomplicated malaria cases, 70% of those with access sought treatment within 3 days across all age groups. For severe malaria cases, 80% of those with access sought treatment within 2 days of symptoms across all age groups [40,41]. Case management was maintained at these rates even after simulating different larviciding strategies. This framework assumed target vectors were susceptible to biolarvicides and parasites to Artemether-Lumefantrine throughout the intervention period (Table 1).

**Table 1.** Summary of model parameters and data sources used in the simulation framework.

| Category | Parameter | Value/ Description | Data Source |
| --- | --- | --- | --- |
| **Model platform** | Simulation model | EMOD v2.20 individual-based model for *P. falciparum* | [36] |
| **Vector ecology & habitats** | Habitat seasonality calibration | Monthly habitat abundance calibrated to field data from rural SE Tanzania | [37] |
|  | Habitat co-sharing | 8% of larval habitats contain both *An. arabiensis* and *An. funestus* | [37] |
| **Larviciding parameters** | Larvicide efficacy & decay | Daily residual decay profile based on bioassay | This study (bioassay described later) |
| **Vector feeding behavior** | Indoor feeding fraction (*An. arabiensis*) | 63.1% | [38] |
|  | Indoor feeding fraction (*An. funestus*) | 78.2% | [38] |
| **ITN characteristics** | Initial killing effect | ~70% in experimental houses | [39] |
|  | Killing effect decay | Exponential decay to ~40% by year 2 | [39] |
|  | Biting-blocking effect | 95% initial; exponential decay over 4 years | Assumption based on [39] |
| **Insecticide resistance** | *An. arabiensis* homozygous resistant | 11% mortality (89% resistance) | Msugupakulya, unpublished |
|  | *An. funestus* homozygous resistant | 2% mortality (98% resistance) | Msugupakulya, unpublished |
| **Treatment behavior & case management** | Access to care | 80% of individuals | [40,41] |
|  | Uncomplicated malaria treatment-seeking | 70% (among those with access) within 3 days | [40,41] |
|  | Severe malaria treatment-seeking | 80% (among those with access) within 2 days | [40,41] |

### Biolarvicides

Biolarviciding was incorporated into the life cycle component of the model to simulate reductions in larval survival and densities. Due to limited prior information on larvicide efficacy and decay, we assumed a daily residual efficacy profile derived from experimental bioassays (described below). Four commercially available biolarvicides, including *Bacillus thuringiensis israelensis* (*Bti*)- and *Bacillus sphaericus* (Bs)-based formulations, were tested at recommended doses against *An. funestus* and *An. arabiensis* larvae, and the observed mortality decay over time was used to parameterize the model.

#### Experiments to assess the bio-efficacy and residual activity of the biolarvicides

The assessment of the decay profile and residual activity of the biolarvicides was done on four different biolarvicides: two granular biolarvicides manufactured by Valent Biosciences (Libertyville, IL 60048 USA) (VectoBac (containing *Bti*) and VectoMax (containing *Bti* and *Bs*) and two liquid biolarvicides manufactured by Tanzania Biotech Products Limited (Kibaha, Tanzania) (Bactivec (containing *Bti*) and Griselesf (containing *Bs* only)). We first created semi-natural habitats, as explained in our previous study [42]. Briefly, we buried 60 basins with a diameter of 41 cm and a height of 14 cm. Inside the basins, we added a small amount of sand and vegetation from natural habitats. We then filled the basins with untreated well water. The habitats were allowed to settle for at least two days to condition. Afterwards, the simulated habitats were then divided into ten groups of six habitats and randomised so that five groups were designated for tests of biolarvicides against *An. funestus* and the other five for *An. arabiensis*. In each habitat, we introduced equal numbers of respective lab-reared larvae of different instars (second to fourth), totalling 100 larvae. For each species (*An. funestus* and *An. arabiensis*), the five groups of habitats were treated as follows: i) Six habitats were treated with 146mg of VectoBac; ii) Six habitats were treated with 146mg of VectoMax; iii) Six habitats were treated with 65μL of Bactivec; iv) Six habitats were treated with 130 μL of Griselesf; and v) Six habitats were not treated with any biolarvicides and served as controls. These dosages were selected based on manufacturers’ recommended doses and a previous study [42].

Each habitat was supplemented with mosquito larvae food (Tetramin fish food) and left for 24 hours while mortality was monitored. We exhaustively counted all live mosquito larvae remaining in the habitat and assumed that the missing larvae had died. After this, we removed all live larvae that had been exposed the previous day and added 100 new larvae of the respective species to each corresponding habitat every 24 hours, and mortality was monitored every 24 hours. We stopped the experiment when mortality in all treatments was equal to that in the controls. This allowed us to obtain the larvicide decay profile for use in the simulations.

### Scenarios simulated

In southeastern Tanzania, an entomological survey conducted a year before this work estimated EIR for malaria to be ∼26 infectious bites per person per year (ib/p/y) [37]. Household ownership of ITNs is very high, reaching up to 99%, though actual net usage at night can be as low as 41% (*Bofu et al., unpublished*). The simulation was designed to achieve an EIR of ∼26 ib/p/y in the second year of the ITNs at coverage of approximately 40%. This EIR was calculated between November and October of the following year. Three different larviciding implementation scenarios were simulated using larvicides lasting less than a week and larvicides lasting for more than a week: i) targeting aquatic habitats of both *An. funestus* and *An. arabiensis* (i.e. broadcast larviciding), ii) a campaign in which habitats containing only *An. arabiensis* were ineligible for larviciding (i.e. *funestus*-targeted larviciding), iii) and a campaign in which habitats containing only *An. funestus* were ineligible for larviciding (i.e. *arabiensis*-targeted larviciding).

Larviciding was simulated for three different durations as a campaign lasting either four months (120 days), five months (150 days), or six months (180 days). All simulations were also run with six different coverage levels of larviciding (i.e., 0%, 40%, 50%, 60%, 70%, and 80%) and two different retreatment intervals for the habitats (i.e., weekly or fortnightly). Additionally, each of these simulations was set to start at a different time to correspond to different seasons in southeastern Tanzania: i) February 1^st^, corresponding to the beginning of the rainy season; ii) April 1st, corresponding to the middle of the rainy season; iii) June 1^st^, corresponding to the transition period between the end of the rainy season and the beginning of the dry season; iv) August 1^st^, corresponding to the middle of the dry season; and v) December 1^st^, corresponding to the end of the dry season (Fig. 1).

**Figure 1.**
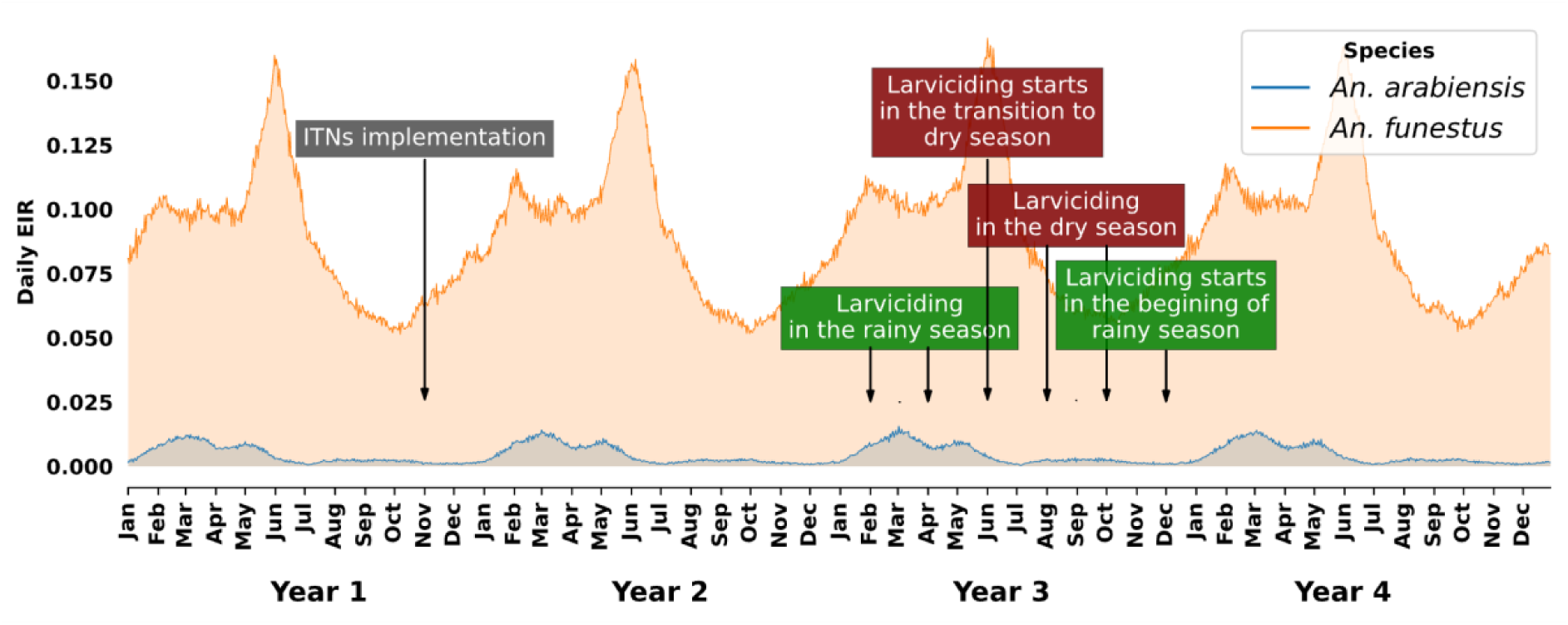
Seasonal fluctuations in malaria transmission. The orange and blue shaded areas and lines depict the distinct patterns of *An. funestus* and *An. arabiensis* between the rainy season (December to May), characterized by peak malaria transmission, and the dry season with lower vector numbers. This figure also illustrates the various start dates and durations of larviciding campaigns incorporated into the simulation. Each campaign was evaluated for one year, starting from the first day of its implementation.

Across all scenarios, ITNs were implemented at three different coverage levels (i.e., 0%, 40%, and 80%). Based on the analysis conducted during the simulation setup, it was observed that ITNs, when implemented alone, would result in the lowest under-five incidence if distributed in November, the transition from dry to rainy season.

Consequently, ITNs were distributed during the same time for these simulations. Twenty stochastic simulations were run for each larviciding campaign.

To evaluate the effectiveness of various larviciding strategies in reducing malaria burden, we compared the relative decrease in clinical cases among children under five and the entomological inoculation rate (EIR). We initially assessed the impact of larvicides alone on malaria incidence in under-fives and EIR, considering factors such as the timing of intervention and the frequency of habitat treatment.

Simulations without any ITNs served as a baseline. In scenarios where we explored the potential of larviciding to supplement ITNs, simulations with 40% ITN coverage and no larviciding distribution were used as a baseline. This approach mirrored the conditions prevalent in rural villages of southeastern Tanzania. We assessed the reduction in EIR and clinical cases among children under five attributable to both increased ITN coverage and various combinations of larviciding with different ITN coverages over different intervention periods. Each campaign scenario was evaluated over one year, starting from the first day of larviciding implementation and concluding 365 days later. This analysis helped to determine whether the combined application of ITNs and larviciding yielded a greater impact than ITNs alone.

### Estimating operational costs of different larviciding strategies

To estimate operational costs for different larviciding strategies, we used aquatic habitat data from a recent field survey in eight villages in south-eastern Tanzania, which documented substantial seasonal and species-specific variation in habitat abundance and size [35]. In these villages, the median number of habitats per village that had either *An. funestus* or *An. arabiensis* was 19 in the dry season and 41 in the rainy season. Median total habitat area was 17,864 m² in the dry season and 18,359 m² in the rainy season; this similarity reflects the inclusion, during the dry-season survey, of more villages, some sampled during the transition from the rainy to the dry season, when residual surface water persisted and formed a small number of unusually large, stable aquatic habitats that disproportionately contributed to total habitat area. Species-specific habitat areas indicated that *An. funestus* habitats were typically fewer but larger (median 7,683–12,309 m²), while *An. arabiensis* habitats were also few but smaller (median 6,228–7,299 m²) and dynamic [35].

Operational costs were calculated using a unit cost of US$0.024 per m² treated, derived from a public provider–perspective costing of microbial larviciding in rural Tanzania [43]. This unit cost reflects recurrent programme implementation expenses, including larvicide procurement, field and supervisory staff time, transportation, training, community sensitization activities, equipment use and maintenance, and routine administrative and communication costs. For each e intervention strategy, we assumed a four-month campaign with fortnightly larvicide applications and 80% coverage of identified aquatic habitats. Separate cost scenarios were developed for interventions targeting An. funestus only, An. arabiensis only, or both species simultaneously. These operational cost estimates were subsequently linked to modeled reductions in entomological inoculation rate (EIR) and malaria incidence to assess the cost-effectiveness of each strategy.

## Results

### Residual efficacy and decay profiles of the biolarvicides

Semi-field trials evaluating two *Bti*-only biolarvicides (BactiVec and VectoBac) and two *Bs*-containing biolarvicides (VectoMax and Griselesf) revealed distinct efficacy profiles. VectoMax and Griselesf had superior performance, achieving an initial efficacy exceeding 98% in reducing larval populations within habitats. Their residual effect persisted for an extended period, with daily efficacy remaining around 50% for at least four days post-treatment and detectable activity lasting up to seven days. In contrast, the *Bti*-only products exhibited more variable efficacy, with initial effects (on the first day of exposure) ranging between 40% and 99% reduction of live larvae. Furthermore, their efficacy waned more rapidly, lasting for a maximum of six days (Fig. 2). Based on these results, VectoMax decay profiles were used to represent biolarvicides with efficacy lasting longer than one week, and VectoBac was selected to represent biolarvicides with shorter residual activity. This choice was informed by the observation that both products showed broadly comparable efficacy and decay patterns across An. funestus and An. arabiensis, with overlapping confidence intervals and no consistent species-specific divergence over time (Fig. 2). The pooled efficacy of these two products over time informed the decay function incorporated into our simulation model (Fig. 3).

**Figure 2.**
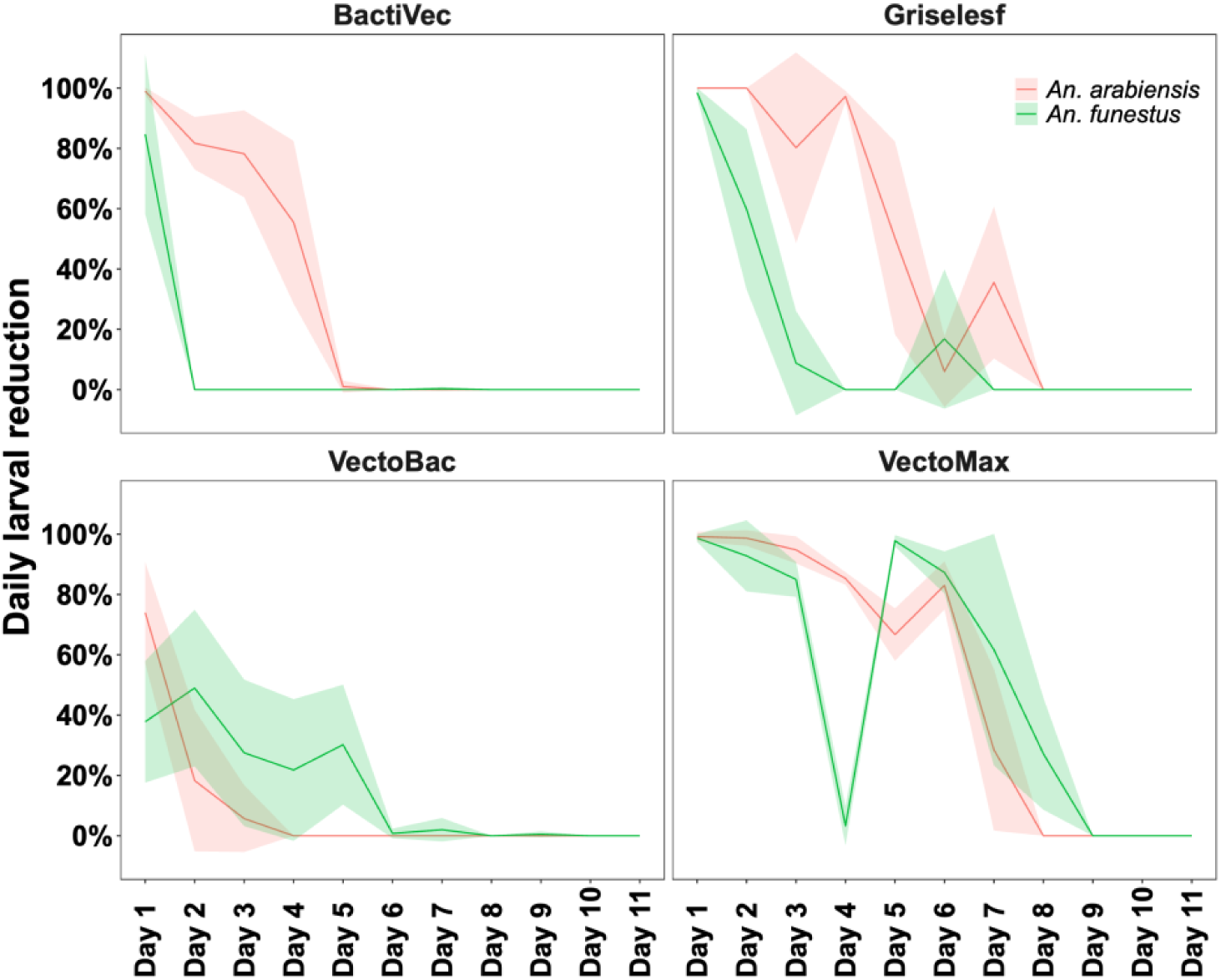
Decay profiles of VectoMax and VectoBac against *An. funestus* and *An. arabiensis* in the simulated habitats.

**Figure 3.**
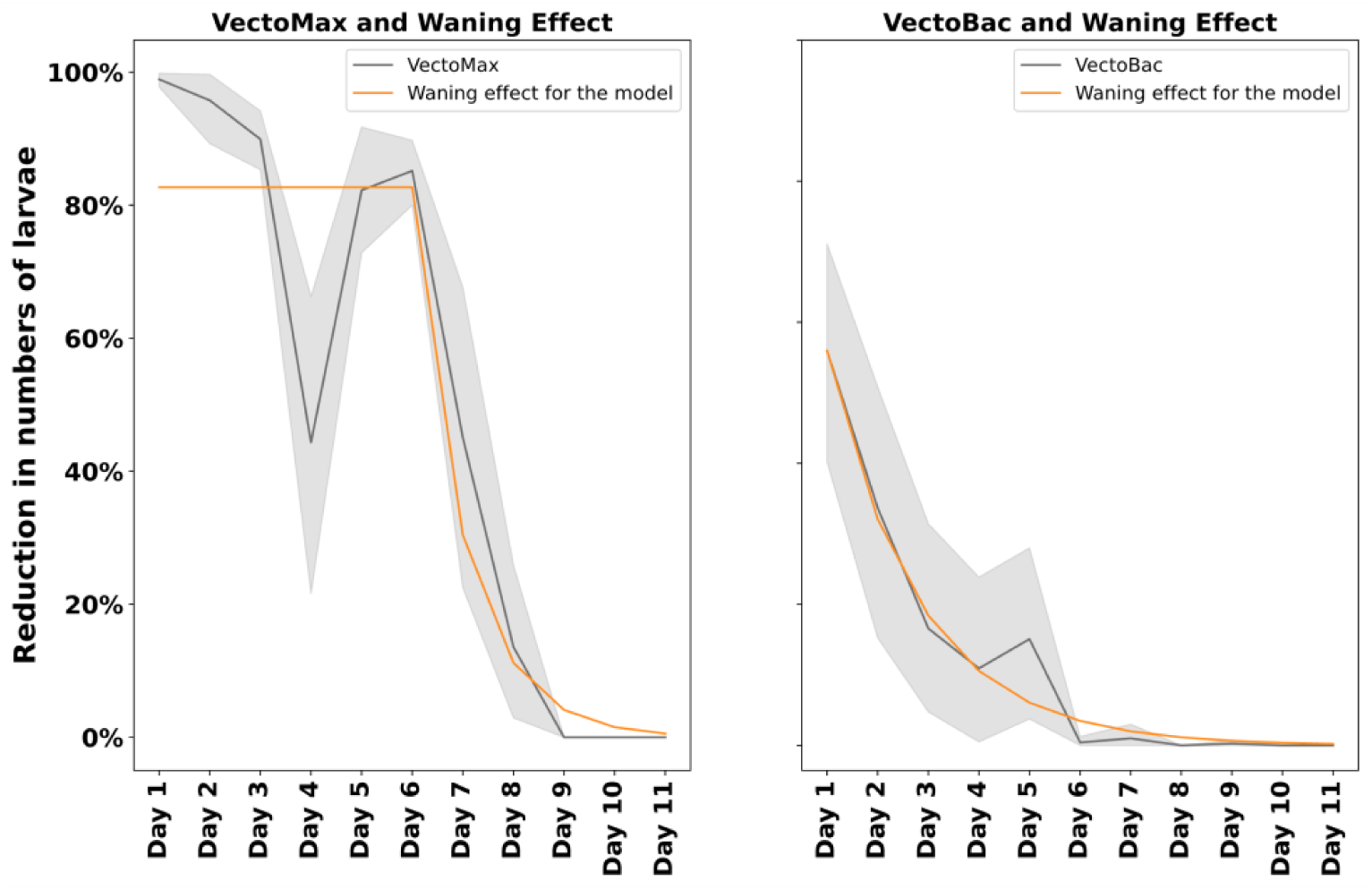
Pooled daily efficacy of VectoMax and VectoBac against *An. funestus* and *An. arabiensis* and the waning effect used in the simulations.

### Impact of different biolarvicides, larviciding strategies, and campaign coverage on malaria transmission and incidence in the absence of ITNs

To assess the impact of different biolarvicides and larviciding strategies on malaria transmission, we conducted an analysis focused solely on larviciding, excluding the use of ITNs. The intervention lasted for four months, beginning in February, just before the onset of the peak malaria transmission season, and continued through the peak period (Fig. 1). The impact was evaluated over a full year from the start of larviciding. In the baseline scenario, which included no vector control intervention, the EIR was approximately 35 ib/p/y, and malaria incidence was about 3.3 clinical cases per child per year (cases/c/y).

Adding moderately long-lasting larvicides (with residual efficacy greater than one week) significantly reduced malaria transmission and cases when targeting either *An. funestus* habitats alone or both *An. funestus* and *An. arabiensis* habitats. Even with less frequent applications (fortnightly) and lower coverage (40% of habitats), these larvicides significantly reduced EIR by over 35% and malaria cases in children under five by more than 16%. In contrast, shorter-acting biolarvicides were far less effective, achieving no more than a 21% reduction in EIR and a 6% reduction in malaria cases among under-fives, even with frequent weekly treatments and 80% larviciding coverage (Fig. 4). As a result, subsequent modelling focused on the potential of moderately long-lasting larvicides. While using moderately long-lasting biolarvicides to control mosquitoes can be very effective, its success depends on a few key factors: which mosquito species are targeted, how much area is covered, and how often treatments are applied.

**Figure 4.**
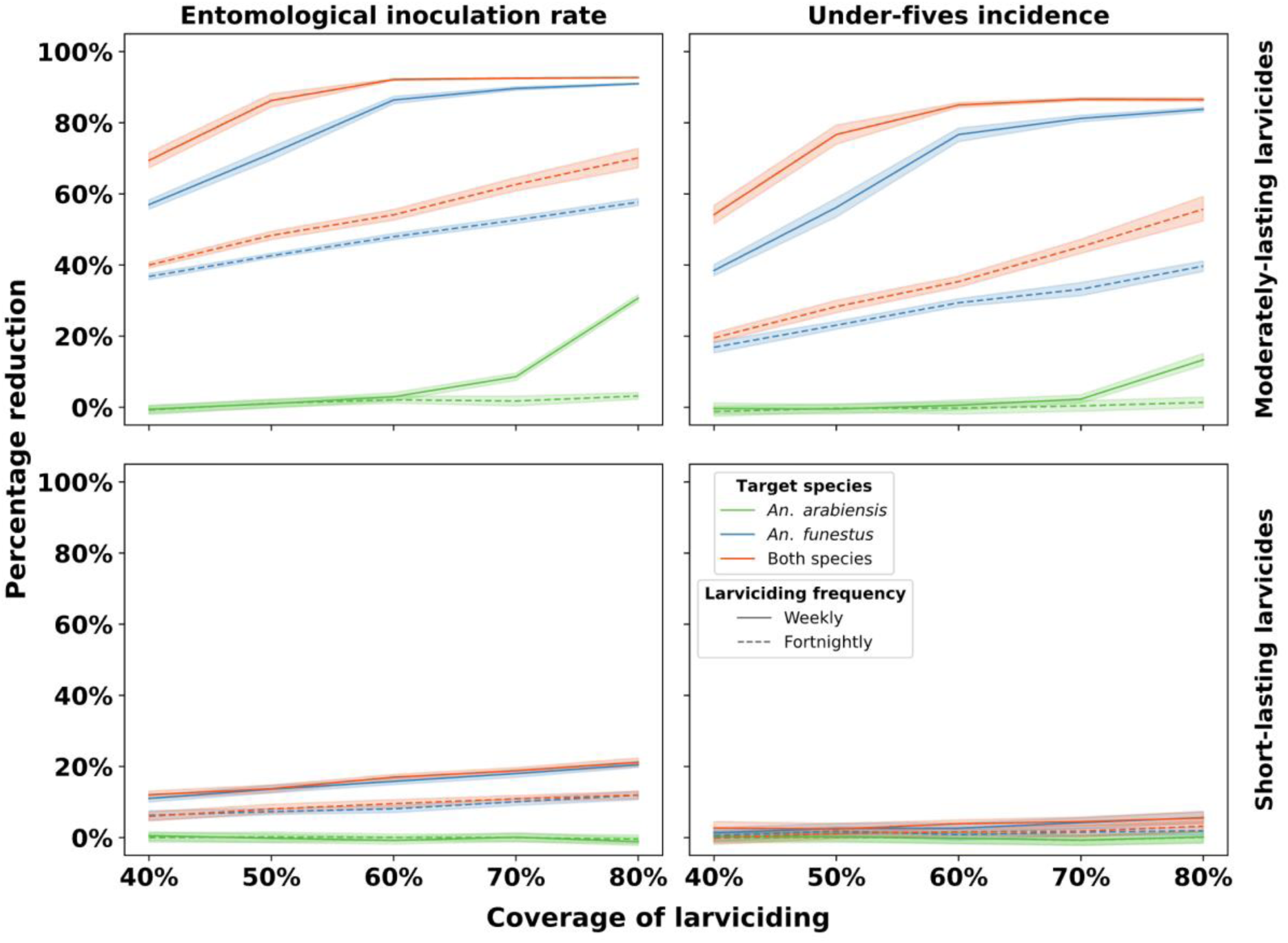
Modeled impact of a larviciding campaign starting in February (before the peak malaria transmission season) on reducing EIR and malaria incidence among under-five children. Larviciding campaign of four months using either moderately longer-lasting or short-lasting biolarvicides, with habitat treatment once a week or once fortnightly. The reduction in EIR and incidence was determined by comparing the larviciding scenario (the only intervention) against a baseline scenario without any intervention. The baseline annual EIR in these simulations was approximately 35 ib/p/y, and the incidence of malaria cases was 3.3 cas/c/y.

**Figure 5.**
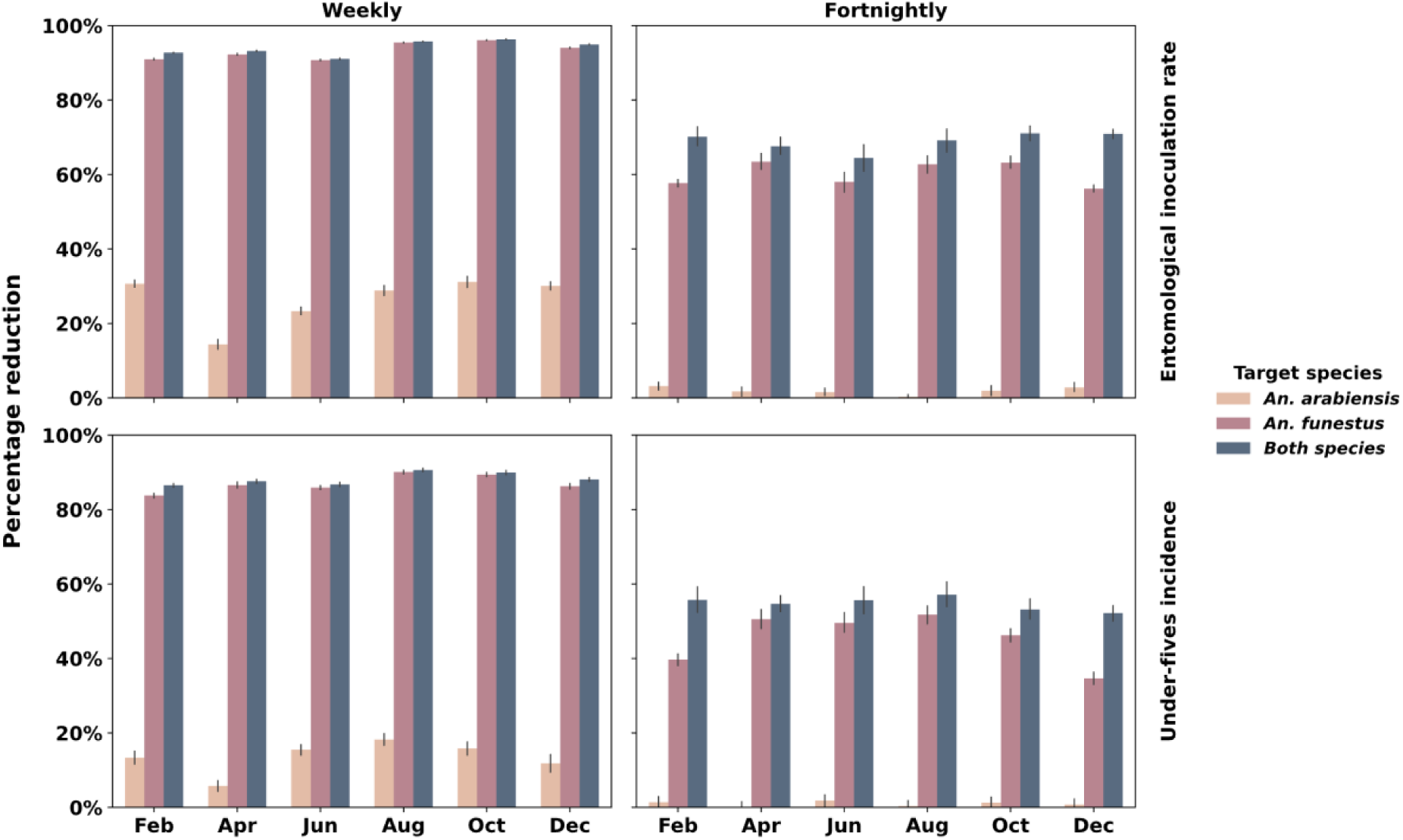
The modeled impact of larviciding interventions starting on different dates on reducing the EIR and malaria incidence among children under five years. The model explores various scenarios with different start dates and frequencies of larviciding, assuming a consistent four-month campaign period and 80% coverage of aquatic habitats. To assess the impact of larviciding, the reduction in EIR and malaria incidence was calculated by comparing the intervention scenarios to a baseline scenario where no larviciding was implemented. The baseline annual EIR in these simulations was approximately 35 ib/p/y, and the incidence of malaria cases was between 3.3 and 3.4 cas/c/y.

In the targeted scenarios using the moderately long-lasting biolarvicides, significant reductions in malaria were observed when larviciding focused on either *An. funestus* alone or both *An. funestus and An. arabiensis*. At 50% larviciding coverage, these strategies achieved approximately 40% reductions in EIR and 20% reductions in malaria incidence among under-five children. However, targeting *An. arabiensis* alone yielded much smaller effects, and even at 80% larviciding coverage, EIR reductions reached only 30%, with a corresponding 13% drop in malaria incidence (Fig. 4).

As expected, the simulations showed that increasing larviciding campaign coverage significantly reduces both EIR and malaria cases. For example, treating aquatic habitats weekly when targeting *An. funestus* alone or both *An. funestus and An. arabiensis* could reduce EIR by 57–69% at 40% larviciding coverage, while the reduction would increase to 91–93% at 80% larviciding coverage. Similarly, malaria cases could decrease by 38–54% at 40% larviciding coverage and by 84–87% at 80% larviciding coverage when targeting *An. funestus* alone or both species. Additionally, reducing the treatment frequency from weekly to fortnightly was predicted to weaken the impact of larviciding. Under fortnightly treatments, EIR reductions fell to 37–40% at 40% larviciding coverage compared to 58–70% EIR reductions at 80% larviciding coverage, when targeting either *An. funestus* alone or both species.

Concurrently, malaria incidence was projected to fall by only 16–19% at 40% larviciding coverage, and by 40–56% at 80% larviciding coverage when both species were targeted (Fig. 4).

### Influence of start date on larviciding efficacy of moderately longer-lasting biolarvicides in the absence of ITNs

Our simulations evaluated the impact of varying larviciding start times across rainy and dry seasons, and different treatment frequencies, on malaria transmission control. The results demonstrated that larviciding can substantially reduce malaria transmission regardless of the season in which the intervention is initiated. This analysis considered a one-year period, during which the baseline annual entomological inoculation rate (EIR) was approximately 35 ib/p/y, and malaria incidence ranged from 3.3 to 3.4 cases/c/y in the absence of vector control. A four-month larviciding campaign with weekly treatments covering 80% of habitats led to a dramatic reduction in annual EIR (between 91% and 96%) and malaria incidence in children under five (between 84% and 91%). These reductions were consistent across campaigns (four-months) starting in different seasons and targeting either *An. funestus* alone or both *An. funestus* and *An. arabiensis*. Campaigns beginning in the dry season were predicted to result in slightly greater decreases in both EIR and incidence compared to those starting in the rainy season. Furthermore, simulations revealed that fortnightly larviciding at the same coverage level resulted in a noticeable decrease in effectiveness. Nevertheless, even with treatments every two weeks, larviciding still achieved substantial reductions, between 57% and 71% in EIR and between 35% and 57% in malaria incidence.

### The potential of larviciding when added to complement ITNs

At baseline, under 40% ITN coverage, the February larviciding campaign began with an EIR of 29 ib/p/y and a malaria incidence rate of 3.2 cas/c/y. In comparison, the August campaign under the same ITN coverage started with a slightly higher EIR of 32.9 ib/p/y and an incidence rate of 3.3 cas/c/y. When ITN coverage was increased to 80%, the February campaign began with a lower EIR of 21.4 ib/p/y and an incidence rate of 2.8 cas/c/y, whereas the August campaign started with a higher EIR of 33.5 ib/p/y and an incidence rate of 3.4 cas/c/y.

Across all scenarios, adding larviciding to ITNs produced substantially greater reductions in transmission than ITNs alone. For campaigns beginning in February, 80% ITN coverage by itself reduced EIR by 26% and malaria cases by 15% relative to baseline (40% ITN coverage). Adding larviciding further increased the impact, with reductions in both EIR and cases rising steadily with larviciding coverage. When *An. funestus* alone or both *An. arabiensis* and *An. funestus* were targeted, the combined interventions reduced EIR by 82–92% and cases by 73–86% relative to baseline. For campaigns started in August, the combined interventions remained effective, producing up to 61–72% reductions in EIR and 49–59% reductions in cases when targeting *An. funestus* or both species (Fig. 6).

**Figure 6.**
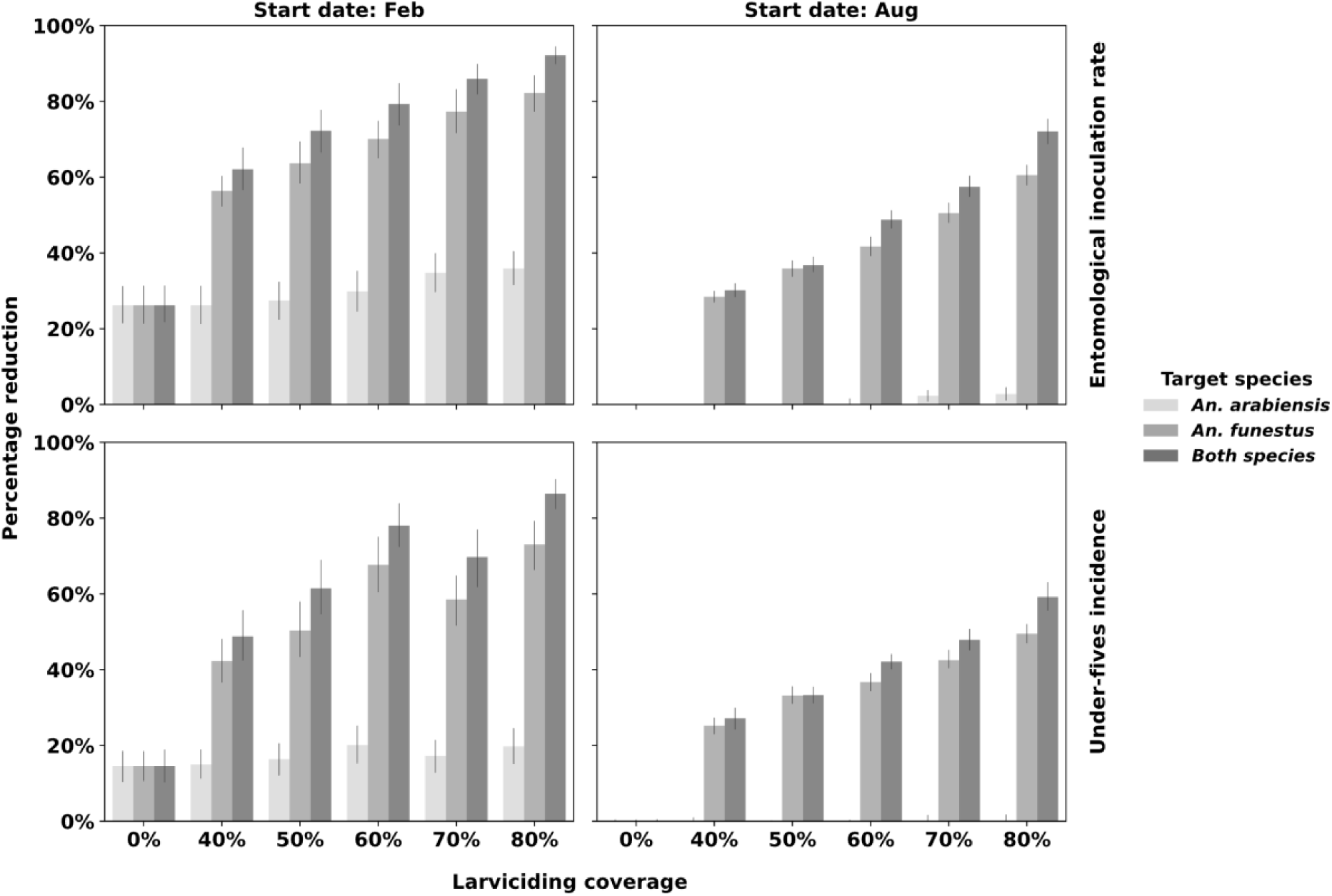
Impact of combined ITN and larviciding interventions on EIR and the incidence of malaria among children under-fives. The analysis considers two different larviciding campaign start times: the rainy season (February) and the dry season (August). Various scenarios were modeled with a campaign duration of four months, with fortnightly habitat larviciding and coverage levels. To assess the impact of larviciding, the reduction in EIR and subsequent malaria incidence was calculated by comparing each intervention scenario to a baseline scenario where only 40% ITN coverage was implemented without any additional interventions.

The analysis of the additional impact attributable specifically to larviciding showed that, in the rainy season, adding larviciding to 40% ITN coverage reduced EIR by 62–76% and cases by 46–63%; with 80% ITNs, reductions increased to 77–90% for EIR and 70–85% for cases when targeting *An. funestus* or both species. In the dry season, larviciding on 40% ITNs reduced EIR by 66–73% and cases by 55–61%, and on 80% ITNs, EIR fell by 61–72% and cases by 50–60% (Supplementary Figure S1).

### Comparative performance of different larviciding campaign strategies complemented with ITNs

The impact of fortnightly larviciding campaigns targeting only *An. funestus* was compared to campaigns targeting both *An. funestus* and *An. arabiensis* across different coverages and durations, assuming approximately 40% ITN coverage. Overall, campaigns targeting both species produced greater reductions in EIR and under-five malaria incidence than those targeting only *An. funestus*, particularly when coverage and duration were similar. Campaigns focusing solely on *An. funestus* could achieve comparable impact if they were either longer or implemented at higher larviciding coverage. For example, a five- or six-month campaign targeting only *An. funestus* was predicted to have the same or greater impact than a four-month campaign targeting both species. Similarly, at equal campaign durations, higher coverage targeting only *An. funestus* (e.g., 80%) achieved greater reductions in transmission than campaigns targeting both species at lower coverages (e.g., 40–60%) (Fig. 7).

**Figure 7.**
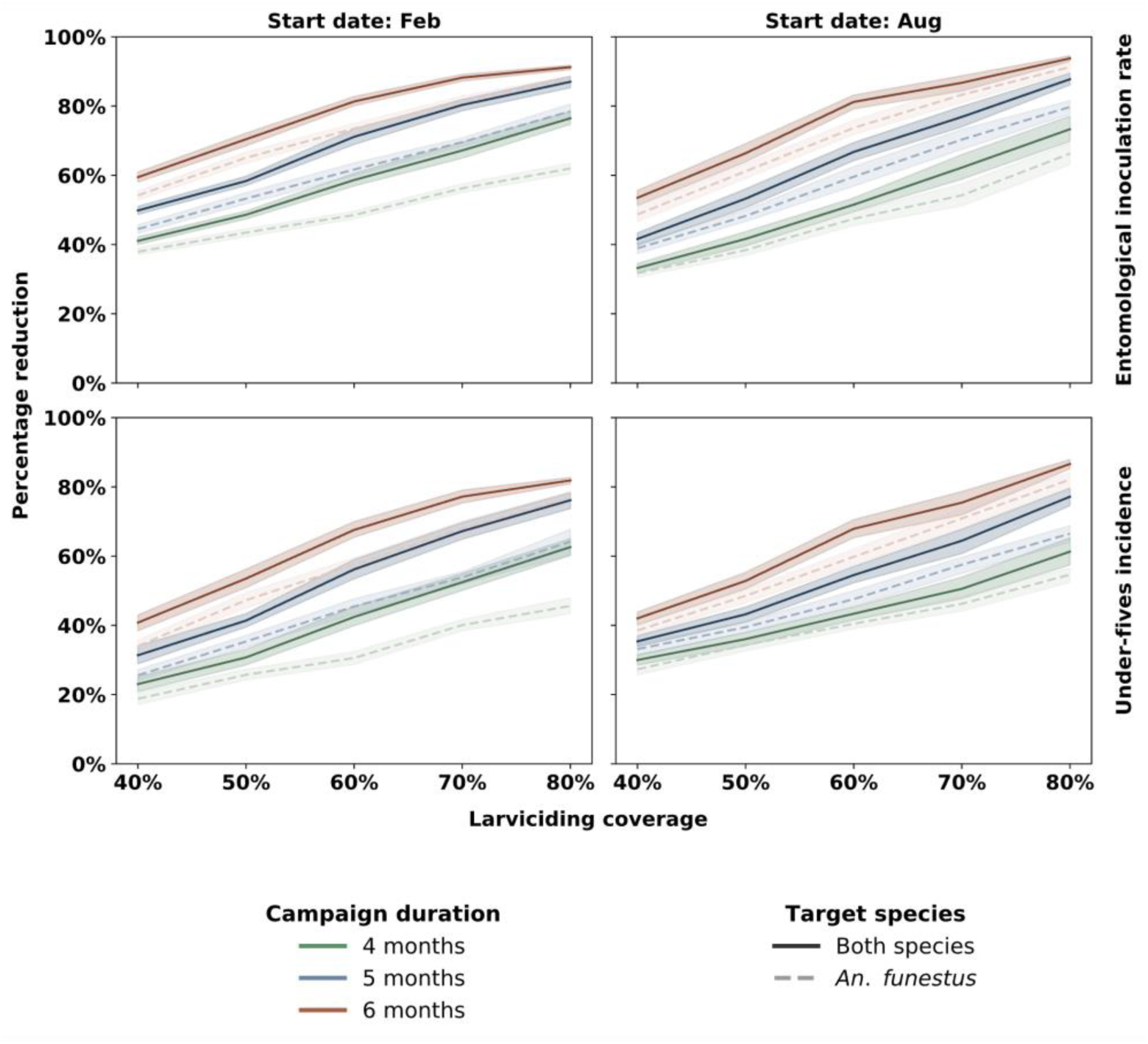
Impact of fortnightly larviciding campaigns on malaria transmission under varying coverage, duration, and vector targets. The figure shows the percentage reduction in EIR and under-five malaria incidence achieved by larviciding campaigns targeting either *An. funestus* only (blue lines) or both *An. funestus* and *An. arabiensis* (red lines) at different larviciding coverages (40% to 80%) and durations (four, five, and six months). Results are stratified by campaign start date (February and August) and assume a constant 40% ITN coverage.

### Operational costs of different larviciding strategies

The estimated median operational cost per village per visit was US$180.8–289.7 when targeting *An. funestus* only, US$146.6–171.8 when targeting *An. arabiensis* only, and US$420.5–432.1 when targeting both species simultaneously_._ When considered alongside modeled epidemiological outcomes, *An. funestus*-targeted larviciding provided the greatest reductions in transmission at the lowest operational cost. At roughly half the cost of combined-species targeting, it achieved substantial reductions in EIR (60–80%) and malaria incidence (50–75%). Expanding treatment to include *An. arabiensis* increased cost by approximately 30–50% but generated only marginal additional epidemiological benefit (less than 15% incremental reduction in EIR and incidence; Fig. 6). Targeting *An. arabiensis* alone was the least efficient option, with costs comparable to the *An. funestus*-only strategy but far smaller reductions in transmission (≤30% EIR reduction; ≤15% incidence reduction), Table 2.

**Table 2.**
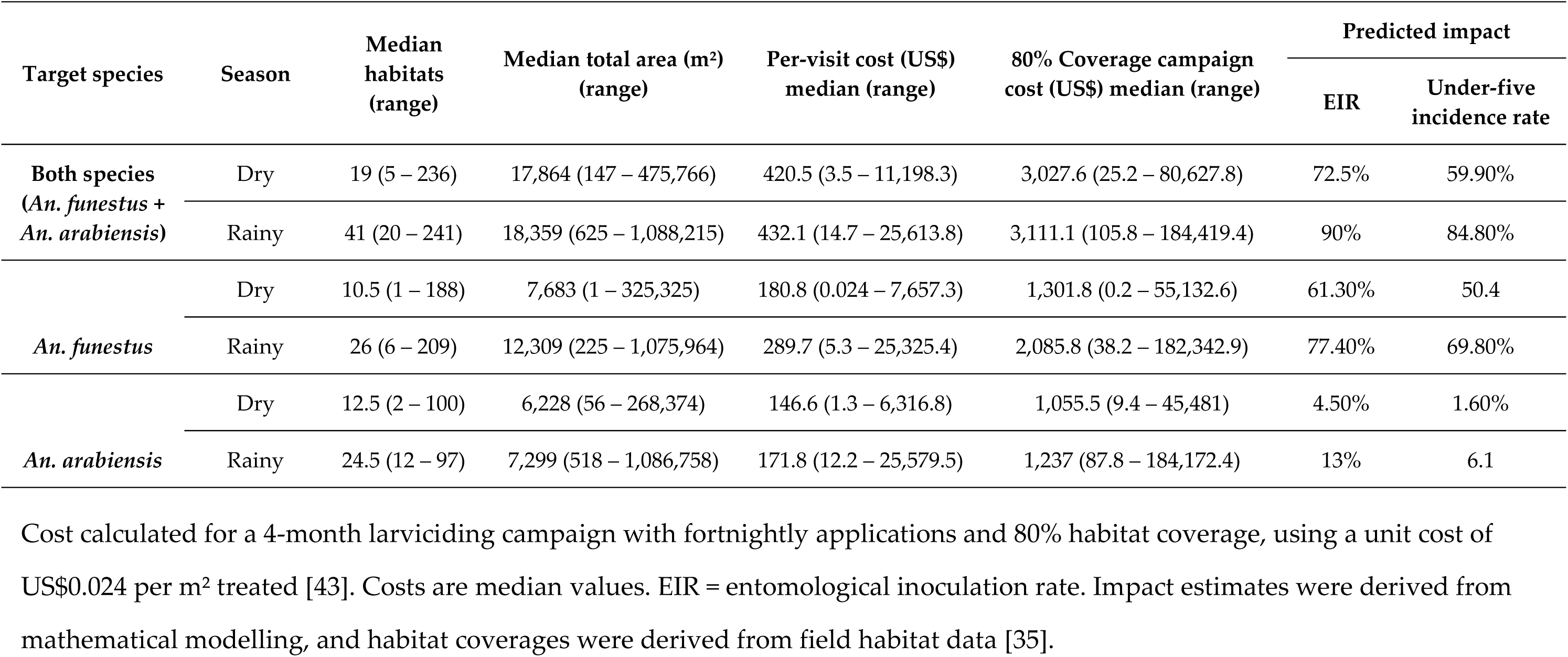
Estimated operational effort and cost of different larviciding campaigns targeting *Anopheles* mosquitoes in south-eastern Tanzanian villages.

| Target species | Season | Median habitats (range) | Median total area (m<sup>2</sup>) (range) | Per-visit cost (US\$) median (range) | 80% Coverage campaign cost (US\$) median (range) | Predicted impact | |
| --- | --- | --- | --- | --- | --- | --- | --- |
|  |  |  |  |  |  | EIR | Under-five incidence rate |
| **Both species**<br>(*An. funestus* + *An. arabiensis*) | Dry | 19 (5 – 236) | 17,864 (147 – 475,766) | 420.5 (3.5 – 11,198.3) | 3,027.6 (25.2 – 80,627.8) | 72.5% | 59.90% |
|  | Rainy | 41 (20 – 241) | 18,359 (625 – 1,088,215) | 432.1 (14.7 – 25,613.8) | 3,111.1 (105.8 – 184,419.4) | 90% | 84.80% |
| *An. funestus* | Dry | 10.5 (1 – 188) | 7,683 (1 – 325,325) | 180.8 (0.024 – 7,657.3) | 1,301.8 (0.2 – 55,132.6) | 61.30% | 50.4 |
|  | Rainy | 26 (6 – 209) | 12,309 (225 – 1,075,964) | 289.7 (5.3 – 25,325.4) | 2,085.8 (38.2 – 182,342.9) | 77.40% | 69.80% |
| *An. arabiensis* | Dry | 12.5 (2 – 100) | 6,228 (56 – 268,374) | 146.6 (1.3 – 6,316.8) | 1,055.5 (9.4 – 45,481) | 4.50% | 1.60% |
|  | Rainy | 24.5 (12 – 97) | 7,299 (518 – 1,086,758) | 171.8 (12.2 – 25,579.5) | 1,237 (87.8 – 184,172.4) | 13% | 6.1 |

A recent field survey in Tanzania revealed substantial seasonal and species-specific variation in aquatic habitat abundance and size [35]. In the dry season, villages had a median of 19 habitats covering 17,864 m² when both *An. funestus* and *An. arabiensis* were present. During the rainy season, the number of habitats increased to 41, and the area covered reached 18,359 m². Disaggregated by species, *An. funestus* habitats were typically fewer but larger (median 7,683–12,309 m²) compared to *An. arabiensis* habitats (median 6,228–7,299 m²).

Applying a unit cost of US$0.024 per m² treated [43], the estimated median campaign cost per village (4 months, 80% coverage, fortnightly treatments) was US$1,301.8-2,085.8 for targeting *An. funestus* only, US$1,055.5–1,237 for targeting *An. arabiensis* only, and US$3,027.6–3,111.1 for targeting both species simultaneously.

When coupled with modeled impacts, *An. funestus*-targeted campaigns deliver substantial additional reductions in EIR (61–71%) and malaria incidence (45–70%) at roughly 30-50% less operational expenditure than the cost of treating both species. Expanding to include *An. arabiensis* increases cost by approximately 30%-50%, but yields only marginal additional epidemiological benefit (less than 15% additional reduction to EIR and incidence rate, respectively, Supplementary Figure S1), while targeting *An. arabiensis* alone is the least efficient, with costs similar to *An. funestus*-targeted campaigns but far smaller transmission reductions (≤30% in EIR; ≤15% in incidence).

Cost calculated for a 4-month larviciding campaign with fortnightly applications and 80% habitat coverage, using a unit cost of US$0.024 per m² treated [43]. Costs are median values. EIR = entomological inoculation rate. Impact estimates were derived from mathematical modelling, and habitat coverages were derived from field habitat data [35].

## Discussion

This study sought to predict the potential impact of LSM using various larviciding strategies and to assess the effectiveness of LSM targeting specific mosquito species on malaria transmission. Our findings indicate that a targeted approach focusing on *An. funestus* alone can achieve nearly the same reductions in malaria transmission as broader campaigns targeting both species, while requiring substantially fewer resources and operational effort. While broad implementation strategies do provide the higher absolute reductions, the additional gains from including *An. arabiensis* are relatively small (<15% extra reduction in EIR and incidence), highlighting critical trade-offs related to how often and how long larvicides are applied and the proportion of habitats they cover. The apparent cost advantage of *An. funestus*–targeted strategies reflect reduced larviciding implementation effort (e.g., area treated and application frequency) and do not include the additional time or resources required to identify species-specific habitats. Therefore, in settings where transmission is dominated by *An. funestus*, targeting this species represents a highly efficient and more achievable strategy, delivering substantial impact potentially at lower cost than multi-species campaigns.

In rural areas, aquatic habitats of *An. arabiensis* tend to vary greatly, and these mosquitoes can be found in small, hard-to-find aquatic habitats. A recent study found that this mosquito can adapt to different types of aquatic habitats based on what is available [35]. This adaptability makes it harder to cover all relevant habitats for both mosquito species effectively, and the challenge of targeting both species using broadcast larviciding may result in less effective outcomes. Since including *An. arabiensis* to larviciding target adds minimal additional impact (<15% reduction) but substantially increases operational costs by 1.5- to 2-fold, focusing solely on *An. funestus* habitats is a far more efficient use of resources. This approach is more practical in the field because *An. funestus* habitats are more limited and predictable in their distribution [33,35]. It also helps to use resources more efficiently, since time and materials that would have been used on less important habitats can instead be used where they are most needed. As a result, targeted campaigns against *An. funestus* can achieve almost the same malaria control outcomes as broad campaigns, while reducing operational costs by roughly 30-50%.

In our study area, rural south-eastern Tanzania, the habitats of *An. funestus*, which tend to be permanent or semi-permanent water bodies, are broadly classifiable as “fixed, few, and findable,” offering a clear operational advantage for targeted larviciding [35]. Field data from this area show that larval surveys identified only 15.2% of all water bodies in the dry season and 16.5% in the rainy season as habitats supporting late-instar *An. funestus* [35]. Although these were relatively few, they made up a large share of the total surface area covered by water bodies, 40.6% during the dry season and 66.4% during the rainy season. Villages had a median of 238 water bodies in the dry season and 342 in the rainy season, meaning that a targeted larviciding campaign would need to survey and treat only around 40 habitats in the dry season and 60 in the rainy season, covering approximately half to two-thirds of the total aquatic surface [35]. This analysis indicates that targeting *An. funestus* habitats in these villages require treating a median area of 7,683–12,309 m² per season, with campaign costs of US$1,301.8-2,085.8 per village (4-month, fortnightly treatments, 80% coverage), compared to US$3,027.6–3,111.1 for campaigns targeting both species. In practical terms, this means only a few hectares per village would need to be treated, which would be manageable within a few days by trained field teams, especially since the habitats are easy to identify and tend to remain in the same locations over time [33,35].

To improve larviciding in diverse sub-Saharan African environments, various propositions and trials have been conducted. Notable among these are mathematical modelling and a field study demonstrating the effectiveness of targeting aquatic habitats based on their capacity to produce adult mosquitoes [23,26]. While this method shows promise, it faces challenges, such as difficulties in identifying productive habitats in the field [27,28]. Efforts to simplify this approach using predictive maps to identify productive habitats have also required specialised skills [23,44]. However, targeting *An. funestus* habitats is inherently simpler because these habitats are fewer, stable, and easily identifiable [1,2], allowing effective interventions without advanced mapping or modeling. Therefore, with the impact of the targeted approach predicted in this study, successful targeted larviciding against *An. funestus* is crucial for optimising malaria control in areas where *An. funestus* is the primary malaria vector, such as in south-eastern Tanzania.

Our study also highlights that initiating larviciding during the dry season can have substantial reductions in malaria transmission. This finding aligns with advocacy for larviciding during the dry season [25]. However, it contradicts previous modelling that suggested a minimal to no impact when larviciding commences during the rainy season [45]. The earlier modelling study assumed a homogeneous vector population of *An. gambiae*, however, in many regions of east and southern Africa, this vector contributes minimally or not at all to malaria transmission during the dry season [15]. In contrast, our predictions incorporated the characteristics of two species: *An. funestus*, which mediates year-round malaria transmission, dominating approximately 95% of malaria transmission, and *An. arabiensis*, which participates primarily during the rainy season, contributing approximately 5% to malaria transmission as noted in a recent study in south-eastern Tanzania [35]. The results of the previous study may hold in areas dominated by *An. gambiae* or *An. arabiensis* that may have little to no transmission during the dry season, such as the Sahel or arid environments. Our predictions are more applicable to regions with *An. funestus* dominating malaria transmission throughout the year. These contrasting predictions from two distinct systems emphasise the crucial need for vector control strategies to be tailored to local vector populations. Failure to consider this can lead to varying impacts. An advantage of implementing larviciding during the dry season is the ease of combining it with other LSM strategies, such as habitat reduction, and it may facilitate achieving higher larviciding coverage.

Several field studies have shown the effectiveness of *Bti*-only biolarvicides in controlling malaria transmission [23,46,47]. However, *Bs*-containing biolarvicides are a compelling alternative, offering longer residual efficacy due to their ability to regenerate within dead larvae [48]. Our simulations predict that biolarvicides combining *Bti and Bs* with a residual efficacy of just one week can achieve greater reductions in malaria transmission compared to *Bti*-only products, even with more frequent application. This highlights the limited value of investing in high-intensity campaigns using short-residual larvicides, reinforcing the strategic advantage of longer-lasting formulations that require fewer treatment rounds. Biolarvicides with substantially longer residual activity than those considered here are already available or under development [49,50]. The use of such longer-acting formulations would likely further amplify the reductions in malaria transmission predicted by our model, potentially allowing for even fewer treatment rounds and lower costs while maintaining or improving epidemiological impact. While *Bs*-containing campaigns may require a higher amount of larvicides applied per square meter in each treatment visit, the overall program cost can be lower due to the reduced frequency of application. This translates to substantial cost savings, particularly in large-scale implementation. Additionally, the moderate and longer-lasting efficacy offers greater flexibility in larvicide application schedules. Delayed residual efficacy and the extended larval development period of *An. funestus* presents a unique advantage for *An. funestus* targeted larviciding as a single application can provide ongoing control for longer periods [42]. Thus, even if aquatic habitat treatments are not administered on a perfectly timed schedule, the residual effect of the larvicide can still significantly impact mosquito larvae that hatched within that timeframe.

There are several limitations of this study. First, while the simulations were based on empirical decay profiles of larvicides, actual field conditions may differ due to factors such as rainfall, water quality, and variability in application fidelity. Second, our model assumes idealized implementation conditions, including perfect detection of mosquito habitats and successful targeting of species-specific habitats, which may not be easily achievable in real-world field operations. While multiple targeting strategies exist, such as prioritizing the most productive habitats, those of dominant vectors, peak transmission periods, low-population nadirs (e.g. dry season), or proximity to human dwellings, our approach focused solely on habitats of the dominant vector (*An. funestus*) as a potentially resource-efficient method yielding outsized impacts, even in rural areas. Additionally, recent advances in mapping technology may facilitate more accurate habitat identification [51–53]. Third, while our results indicate strong impact potential under various scenarios, they are based on modelling outputs; therefore, real-world validation in southeastern Tanzania and other settings where transmission is dominated by *An. funestus* is essential to confirm these projections.

## Conclusion

This study demonstrates that larviciding can substantially reduce malaria transmission in south-eastern Tanzania when implemented at sufficient coverage, frequency, and duration. While targeting both *An. funestus* and *An. arabiensis* offers broad suppression across vector species; our findings show that the additional epidemiological benefit gained by including *An. arabiensis* is relatively small compared to the major increase in operational effort and cost required. In this setting, where *An. funestus* consistently dominates transmission, larviciding campaigns focused solely on *An. funestus* achieves reductions in EIR and clinical incidence that are nearly equivalent to those achieved by treating both species, while likely incurring roughly 30–50% lower costs. Moreover, achieving higher larviciding coverage of *funestus*-targeted campaigns (e.g., 80%) resulted in greater impacts than campaigns targeting both species but achieving lower coverages (e.g., 40%, 50%, or 60%). The restricted distribution, stability, and predictability of *An. funestus* habitats further enhance the practicality of this targeted approach. Given these considerations, prioritizing *An. funestus*-focused larviciding represents a highly efficient strategy for programs operating under resource constraints, with multi-species campaigns offering only marginal additional impact for substantially greater investment.

## List of abbreviations

Bs: Bacillus sphaericus
Bti: Bacillus thrungiensis var israeliensis
COVID-19: Coronavirus Disease of 2019 EIR Entomological Inoculation Rate
EMOD: Epidemiological MODeling software ib/p/y Infectious Bites per Person per Year
cas/c/y: Clinical cases per Under-five Child per Year IRS Indoor Residual Spraying
ITN: Insecticide Treated Nets
LSM: Larval Source Management
WHO: World Health Organization

## Data Availability

All data supporting the main conclusions of this article are included within the article.

## Acknowledgements

The authors would like to thank the Institute for Disease Modeling for providing access to the EMOD platform, which was used to perform the simulations in this study.

## Authors’ contributions

BJM, FOO, ALW, and PS jointly conceived the study and contributed to the design of the research framework. BJM conducted the simulations with guidance and support from PS. BJM drafted the initial manuscript and incorporated critical revisions and feedback provided by FOO, ALW, and PS. All authors reviewed and approved the final version of the manuscript.

## Ethics approval and consent to participate

Ethics approvals for this study were obtained from the Institutional Review Board of Ifakara Health Institute (Ref no: IHI/IRB/No: 32-2021) and the Medical Research Coordinating Committee (MRCC) at the National Institute for Medical Research (Ref no: NIMR/HQ/R.8a/Vol. IX/3761).

## Consent for publication

Permission to publish was granted by The National Institute for Medical Research (NIMR), Tanzania, Ref No. BD.242/437/01C/187.

